# Analytical performance of a delayed-centrifugation blood collection tube for remote blood biomarker assessment

**DOI:** 10.64898/2026.10.05.26364780

**Authors:** You-Rim Lee, Xuemei Zeng, Marissa F. Farinas, Brian J. Lopresti, Rima F. Habte, Kristine A. Wilckens, Thomas K. Karikari

## Abstract

Current biobanking guidelines require in-person blood collection and immediate processing for Alzheimer’s disease (AD) biomarker assessment. We recently reported that the P100 blood collection tubes containing protease inhibitors improve protein biomarker stability over delayed centrifugation periods. However, whether this comparability extends beyond a small set of biomarkers to the broad panels enabled by high-plex platforms has not been established. Here, we performed a head-to-head comparison of 127 neurodegeneration-related biomarkers in paired P100 vs. standard ethylenediaminetetraacetic acid (EDTA) plasma samples from *n* = 43 community-dwelling older adult participants using the NUcleic acid-Linked Immuno-Sandwich Assay (NULISA) proteomic technology. The NULISA assay demonstrated excellent technical reproducibility (intra- and inter-plate coefficients of variation [CVs] <10%). Biomarker levels in P100 and EDTA tubes were strongly correlated, with mean and median Spearman coefficients exceeding ρ = 0.8. Six conventional AD biomarkers showed strong P100–EDTA correlations (ρ > 0.8) with minimal mean differences and no significant proportional bias in Bland–Altman analysis, including plasma Aβ42, GFAP, MAPT, NEFL, p-tau181, and p-tau231, while p-tau217 showed a P100–EDTA correlation of ρ = 0.57. Two biomarker ratios (Aβ42/Aβ40 and p-tau217/Aβ42) also showed minimal bias with no proportional bias detected. Receiver operating characteristic analysis using amyloid status, assessed with positron emission tomography in a subset of participants, showed comparable classification performance between the two tube types, with classification concordance up to 92% and no significant DeLong differences for most (80%) biomarkers. These findings indicate that, on the NULISA platform, P100 tubes yield analytical and classification performance comparable to conventional EDTA tubes, supporting their use for blood-based neurodegeneration biomarker research where immediate sample processing is not feasible.

## Introduction

Blood-based biomarkers for Alzheimer’s disease (AD) have emerged as promising tools for diagnosis and triaging, with active research underway to enable their clinical implementation^1–3^. The field is rapidly advancing in this direction, spurred on by the considerable advantages of blood-based biomarkers over traditional cerebrospinal fluid (CSF) and positron emission tomography (PET) imaging approaches in terms of accessibility, cost-effectiveness, and scalability^4,5^.

A critical prerequisite for reliable blood-based diagnostics is ensuring the pre-analytical stability of protein biomarkers^6^. Current standard protocols emphasize immediate temperature-controlled storage of whole blood, followed by plasma processing within 1-2 hours, requiring reliable electricity supply and centrifugation capacity^7–9^. However, these stringent requirements pose significant challenges in various clinical settings, particularly in rural areas or regions with limited laboratory infrastructure, where samples may require extended transport times before processing^3^.

To address these pre-analytical challenges, blood collection tubes containing protease inhibitors have been developed^10^. Proteolytic degradation represents a primary factor contributing to blood biomarker instability^11,12^. The BD™ P100 tube, coated with a specialized protease inhibitor cocktail, has shown promise in stabilizing plasma biomarkers at room temperature^13,14^. In our recent study, we demonstrated that P100 tubes effectively maintained the stability of several AD biomarkers measured with Single molecule array (Simoa) under room temperature conditions relative to traditional ethylenediaminetetraacetic acid (EDTA) tubes^14^. These include amyloid beta 40 (Aβ40), amyloid beta 42 (Aβ42), Aβ42/Aβ40 ratio, glial fibrillary acidic protein (GFAP), neurofilament light polypeptide (NEFL), and tau phosphorylated at threonine 181 (p-tau181). However, that assessment was limited to a small set of biomarkers measured individually on a targeted platform. Whether P100 tubes perform comparably to EDTA across a substantially broader biomarker panel—and on the higher-plex platforms increasingly used for such panels— remains unknown.

Recent technological advances have enabled highly sensitive multiplex protein quantification. The NUcleic acid-Linked Immuno-Sandwich Assay (NULISA) platform represents a cutting-edge technology capable of quantifying hundreds of purposefully selected protein biomarkers from as little as 10 μL of plasma with attomolar-level sensitivity^15^. This high-throughput capability makes NULISA particularly well-suited for comprehensive analytical validation studies. In contrast to targeted single-molecule immunoassays such as Simoa, which measure a limited number of biomarkers individually, NULISA profiles a far larger and more diverse panel in parallel. Tube-related effects characterized for a handful of biomarkers cannot be assumed to generalize across the full range of biomarkers, leaving many other clinically promising biomarkers unexamined. A broad evaluation is therefore needed to establish the groundwork required to support subsequent validation efforts. Characterizing how P100 tubes perform relative to conventional EDTA tubes on the NULISA platform provides this groundwork, supporting wider adoption of remote blood sampling across a broad range of AD biomarkers in research and clinical settings.

The present study aimed to evaluate the analytical performance of the P100 tubes relative to standard EDTA tubes using the NULISA CNS Disease Panel, which at the time of this study quantified a defined list of 127 biomarkers (Supplementary Table 1). Specifically, we assessed: (1) assay reproducibility; (2) biomarker detectability; and (3) analytical and concordance in amyloid status classification between P100 and conventional EDTA plasma.

## Methods

### Participants

Blood samples were collected from participants enrolled in the Alzheimer’s Pathways Sleep Study (ALPS) cohort within the University of Pittsburgh School of Medicine Department of Psychiatry. The ALPS study, overseen by the Principal Investigator (K. A.W.), is a clinical trial intervention for which the primary aim is to test the effects of a sleep intervention on cognitive function and biomarkers related to Alzheimer’s disease^16^, including carbon-11 labeled Pittsburgh Compound-B ([^11^C]PiB) PET imaging to assess brain Aβ status (A+/A-) and load^17^. All study procedures were approved by the University of Pittsburgh Institutional Review Board (initial approval June 1, 2021; most recent approval February 17, 2026), and all participants provided written informed consent.

Participants were adults aged 65–85 years with no diagnosis of Alzheimer’s disease who self-reported poor sleep quality, confirmed by sleep diary and actigraphy (sleep efficiency ≤ 92%; wake after sleep onset > 20 minutes). They were recruited primarily from the Greater Pittsburgh and broader western Pennsylvania area through community advertisements, participant registries, and referrals. Exclusion criteria, applied through remote screening and medical evaluation, were primarily intended to minimize safety risks related to the intervention or the risk of excessive daytime sleepiness. Participants were excluded for chronic conditions that could not be improved by behavioral time-in-bed restriction or could exacerbate daytime sleepiness, including severe obstructive sleep apnea (assessed by a one-night at-home ApneaLink study), severe psychiatric conditions, and a history of CNS disease or traumatic brain injury. Sleep-affecting medications (antidepressants, antipsychotics, anticonvulsants, and steroids) and routines (night-shift work, excessive caffeine or alcohol use) were also exclusionary, as was employment as a commercial driver, to minimize the risk of sleepiness-related accidents.

In total, 43 participants (62.8% female; median age 69.5 years) were included in the present analysis. Participants contributed samples at up to two time points (T1 and T2), yielding 75 P100 and 71 EDTA samples, of which 69 formed paired P100–EDTA samples (39 at T1 and 30 at T2). Analyses requiring paired data (Spearman correlation and Bland–Altman) used the 69 paired samples, whereas the volcano plot, ROC, and DeLong analyses used all available samples in each group. Participant characteristics, including demographics and [^11^C]PiB PET status, are summarized in Table 1.

**Table 1.** Demographic and clinical characteristics of study participants*.

| Variable | Number of participants |
| --- | --- |
| Number of participants | 43 |
| Sex |  |
| Male (%) | 16 (37.2%) |
| Female (%) | 27 (62.8%) |
| Age (years) | 69.5 (67.4 – 73.8) |
| Education (years) | 16.0 (14.3 – 18.0) |
| Self-identified racial identity |  |
| Non-Hispanic White (%) | 38 (90.5%) |
| Black/African American (%) | 3 (7.1%) |
| Asian Pacific (%) | 1 (2.4%) |
| *APOE* ε4 carriership |  |
| Yes | 8 (21.6%) |
| No | 29 (78.4%) |
| Amyloid beta PET |  |
| Positive (%) | 8 (19.5%) |
| Negative (%) | 33 (80.5%) |

\*Continuous variables are presented as median (1st–3rd quartile). Categorical variables are presented as counts and percentages. Counts and percentages for each item were calculated based on non-missing data. Data were missing for race ( $n = 1$ ), *APOE* ε4 ( $n = 6$ ), and Amyloid beta PET ( $n = 2$ ). *APOE*, apolipoprotein E; PET, positron emission tomography.

Amyloid PET status was determined by expert visual read. The four 5-minute acquisition frames of [¹¹C] PiB PET images were summed into a single image corresponding to 50–70 minutes post-injection. The summed images were smoothed with a 5-mm Gaussian kernel and presented to an expert rater (B.J.L.) for visual interpretation of PiB status using PMOD software (v4.5, Bruker Corp., Billerica, MA, USA). A four-point rating scale permitting equivocation was used (0 = unequivocally negative, 1 = probably negative, 2 = probably positive, 3 = unequivocally positive). Scans rated 0–1 were classified as amyloid-negative and those rated 2–3 as amyloid-positive^16^.

### Blood sample collection and preparation

Blood samples for each participant were collected in the laboratory in the morning for each participant within an hour of their wake time at each of two time points, separated by a 4-week behavioral sleep intervention. All available samples from the two time points were included in the current analysis. For each participant, venous blood was collected into paired tubes: 10 mL EDTA tubes (BD Biosciences, Franklin Lakes, NJ, USA; Cat. No. 366643) and 8.5 mL P100 tubes containing protease inhibitor cocktail (BD Biosciences; Cat. No. 366448). Immediately following blood draw, tubes were inverted 8-10 times to ensure proper mixing and centrifuged at 2,000×g for 15 minutes at 4 °C to separate plasma from cellular material. Plasma samples were aliquoted and stored at −80 °C until NULISA analysis.

### Biomarker quantification using NULISAseq

Plasma samples were analyzed using the NULISAseq CNS Disease Panel on the ARGO HT System (Alamar Biosciences, Fremont, CA, USA), which quantifies 127 neurodegeneration-related protein biomarkers. Prior to analysis, plasma samples were thawed and centrifuged at 10,000 ×g for 10 minutes to remove particulates.

A total of 30 μL of plasma per sample was used for the assay, as previously described^18^. Briefly, samples were incubated with paired capture and detection antibodies conjugated with target-specific DNA barcodes. Following magnetic bead-based purification and ligation, DNA reporter molecules were quantified by next-generation sequencing. Samples were randomized across two assay plates.

Data were normalized using internal controls and inter-plate controls, then rescaled and log2-transformed to generate NULISA Protein Quantification (NPQ) units. The limit of detection (LOD) for each target was calculated as the mean plus three standard deviations of negative control samples on each plate, followed by log2-transformation. The target names and protein names of all 127 biomarkers are listed in Supplementary Table 1.

### Statistical analysis

Statistical analyses were performed using GraphPad Prism version 10 (GraphPad Software, San Diego, CA, USA) and R version 4.4.2. Analyses requiring paired data—Spearman correlation and Bland–Altman analysis—were performed on the 69 paired P100–EDTA samples, whereas the volcano plot, ROC, and DeLong analyses used all available samples in each group (75 P100 and 71 EDTA). Spearman correlation coefficients were calculated to assess the correlation between P100 and EDTA measurements for each biomarker.

For volcano plot analysis, differential abundance— the magnitude of between-tube differences in each biomarker’s levels—was evaluated using the Wilcoxon rank-sum test for each biomarker, with *p*-values adjusted for multiple comparisons using the Benjamini-Hochberg false discovery rate (FDR) method. The rank-sum (rather than signed-rank) test was used because the number of available samples differed between tube types, precluding a paired comparison.

Bland-Altman analysis was performed to assess systematic bias and proportional bias between P100 and EDTA measurements. To assess proportional bias, linear regression was performed with the difference (P100 - EDTA) as the dependent variable and the mean of P100 and EDTA as the independent variable; a significant slope (*p* < 0.05) was considered indicative of proportional bias.

Receiver operating characteristic (ROC) curve analysis was conducted to evaluate classification performance, with optimal cutoff values determined using the Youden’s index. DeLong test was used to compare AUC values between EDTA and P100. Biomarker ratios (Aβ42/Aβ40 and p-tau217/Aβ42) were calculated by subtracting log2-transformed NPQ values.

NULISA assay reproducibility was evaluated using intra-plate and inter-plate coefficients of variation (CVs) calculated for each of the 127 targets using the built-in sample controls included on every NULISA plate by the manufacturer.

Detectability was assessed at two levels: target-level detectability was defined as the percentage of samples with measurements above the LOD for each target, and sample-level detectability was defined as the percentage of targets with measurements above the LOD for each sample. A significance level of *p* < 0.05 was used for all statistical tests.

## Results

### NULISA assay reproducibility

Samples were analyzed across two independent assay plates. Intra- and inter-plate CVs were calculated for each of the 127 biomarker targets, and their distributions were visualized as histograms (Figure 1a-d). Both mean and median CVs for intra- and inter-plate comparisons were within 10%, indicating high assay reproducibility. However, in the overall intra-plate analysis (Figure 1c), six targets exhibited CVs exceeding 20% (NPTX1, MME, PSEN1, SOD1, p-tau217 and Aβ38), indicating lower measurement reproducibility for these targets. In the inter-plate analysis (Figure 1d), eight targets exceeded this threshold (PSEN1, p-tau217, MME, NPTX1, Aβ40, FLT1, Aβ38 and SOD1).

**Figure 1.**
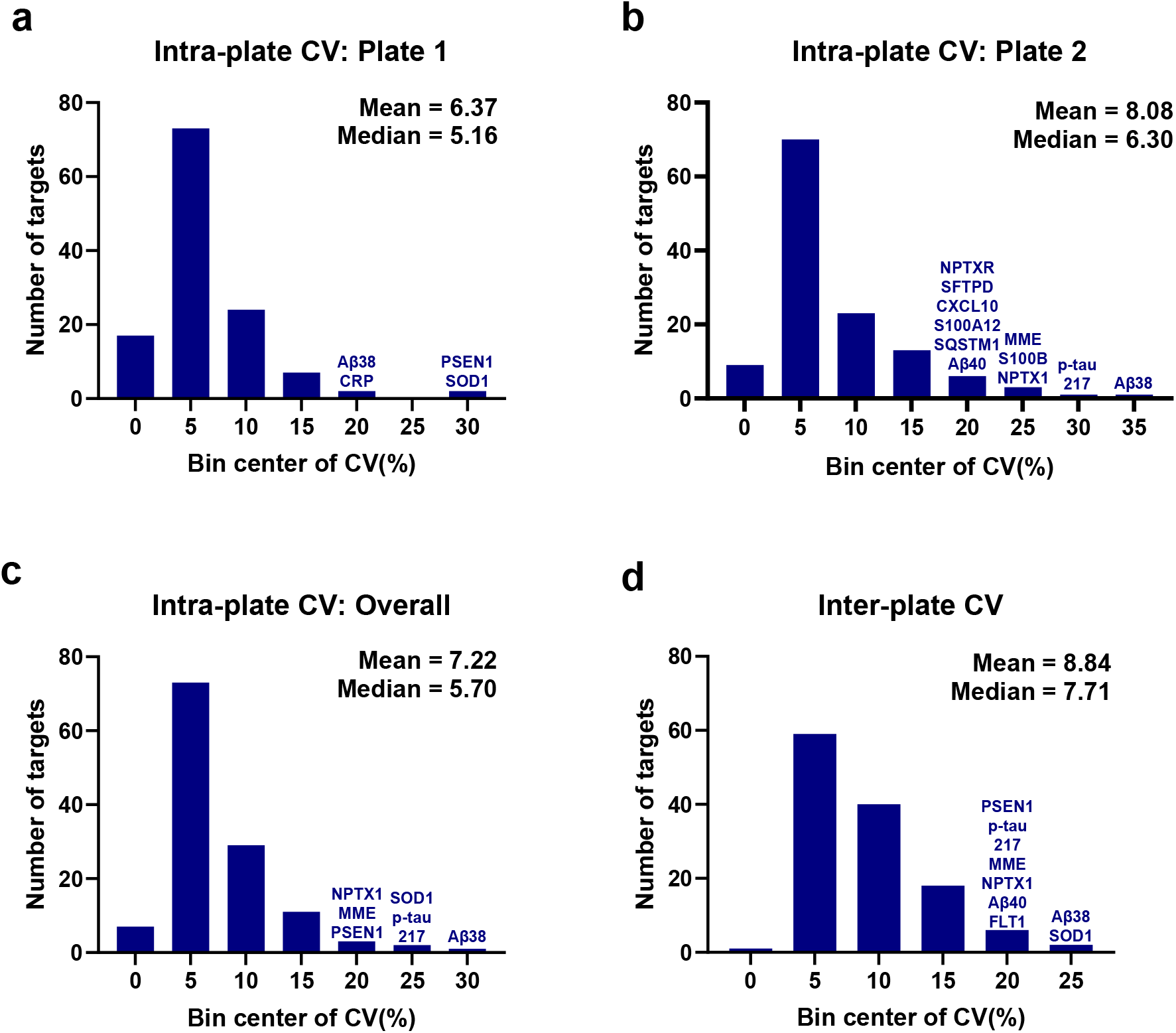
NULISA assay reproducibility. Percentage coefficients of variation (CV) were calculated for intra-plate and inter-plate results across 127 biomarkers using sample controls. Samples were analyzed across two independent assay plates. (a-c) Intra-plate CV distribution for plate 1 (a), plate 2 (b), and combined results from both plates (c). (d) Inter-plate CV distribution. The mean and median CV values for panels (a-d) were within 10%, indicating high reproducibility. For CV bin centers ≥20%, biomarker target names are indicated above the corresponding bars. Aβ40, amyloid beta 40; p-tau217, tau phosphorylated at threonine 217. Full names of all other protein targets are provided in the Abbreviations list and Supplementary Table 1.

### Sample-level and target-level detectability

Sample-level detectability was significantly different between groups: the EDTA group had a mean detectability of 95.9% compared with 94.7% for the P100 group (*p* < 0.0001; Figure 2a). Although this difference reached statistical significance, the absolute magnitude was small (1.17 percentage points). Overall, P100 samples exhibited slightly lower detectability compared to EDTA. Comparison of eight conventional AD biomarkers showed no statistically significant detectability differences for seven targets; NEFL was the exception, exhibiting a significant difference (*p* = 0.026; Figure 2b). Most biomarkers showed NPQ-LOD values well above zero, indicating robust detection, although plasma p-tau217 showed values near the detection limit in both tube types.

**Figure 2.**
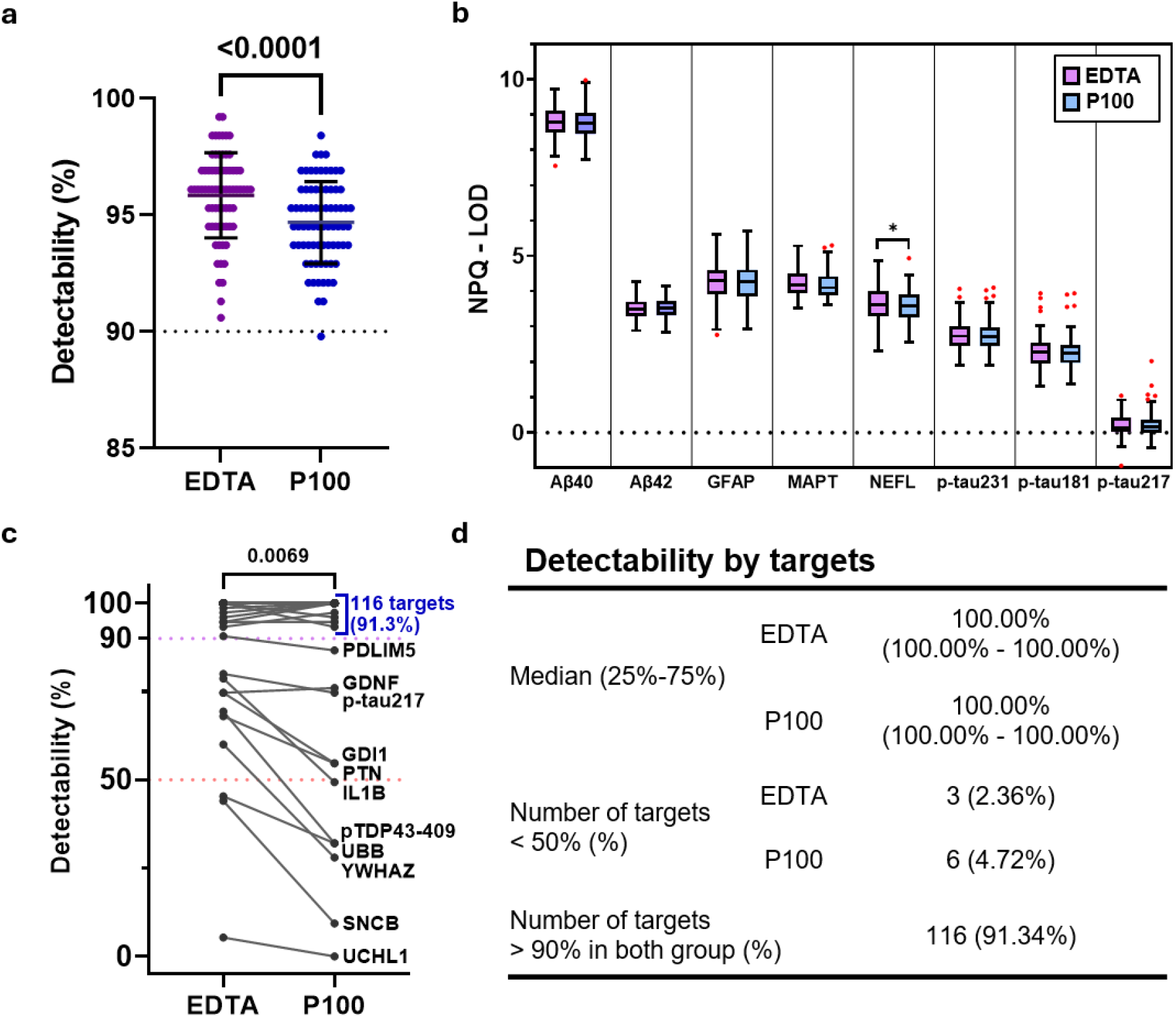
Sample-level and target-level detectability in EDTA and P100. (a)Sample-level detectability comparison between EDTA and P100 groups. Each dot represents a single sample, with the y-axis showing the average detectability across all 127 targets. Both groups exhibited median detectability values > 94%. Error bars represent standard deviation. P-value is indicated on the graph. (b) Comparison of eight conventional AD biomarkers between EDTA (left, purple) and P100 (right, blue) groups using Tukey’s box-whisker plots. The y-axis shows NPQ minus LOD; values above zero indicate detection above the limit of detection. Red dots indicate outliers. \**p* < 0.05. (c) Target-level detectability line plot comparing EDTA and P100. Of 127 targets, 116 (91.3%) showed > 90% detectability in both groups. Targets with < 90% detectability are labeled; all except GDNF showed decreased detectability in P100. (d) Summary table of detectability metrics by target, comparing EDTA and P100 groups. Aβ40, amyloid beta 40; Aβ42, amyloid beta 42; GFAP, glial fibrillary acidic protein; MAPT, microtubule-associated protein tau; NEFL, neurofilament light polypeptide; p-tau181, tau phosphorylated at threonine 181; p-tau217, tau phosphorylated at threonine 217; p-tau231, tau phosphorylated at threonine 231. Full names of all other protein targets are provided in the Abbreviations list and Supplementary Table 1.

For target-level detectability, median values and interquartile ranges were 100% for both groups, and 116 targets (91.3%) had detectability exceeding 90% in both P100 and EDTA samples (Figure 2c-d). A target was considered “detectable” if its detectability exceeded 50% of samples. Six target analytes fell below this threshold in the P100 group, compared with three in the EDTA group (Figure 2d). Notably, all three low-detectability targets in the EDTA group (SNCB, UCHL1, and UBB) were also among the six low-detectability targets in P100, with three additional targets (YWHAZ, pTDP43-409, and IL1B) showing reduced detectability only in P100. The distribution of NPQ-LOD values across all 127 targets for both EDTA and P100 groups is presented in Supplementary Figure 1. Overall, EDTA and P100 showed similar patterns and comparable signal levels for the vast majority of biomarkers.

### Comparison between P100 and EDTA plasma

We examined the biomarker levels between P100 and EDTA plasma samples. Volcano plot analysis revealed that, relative to EDTA, the P100 group had significantly lower concentrations for 33 targets and significantly higher concentration for 1 target (hemoglobin subunit alpha, HBA1) (Figure 3a). Correlation analysis across all targets showed a mean and median Spearman correlation coefficient above 0.8 (Figure 3b), with 77 targets demonstrating strong correlation (ρ ≥ 0.8) and 111 targets showing moderate to strong correlation (ρ ≥ 0.6). The top 10 targets with the highest correlations were identified, with SAA1 achieving the highest value (ρ = 0.996; Figure 3c). Nine targets showed weak correlation (ρ < 0.6), including GDI1, HBA1, and SNAP25; HBA1 exhibited the lowest correlation (ρ = 0.15), corresponding to the largest fold-change observed in the volcano plot. To examine whether low correlation was associated with signal intensity or detectability, we analyzed the relationship between correlation coefficients and mean NPQ values (ρ = 0.190, *p* = 0.033) and detectability (ρ = 0.220, *p* = 0.013). Both showed weak but statistically significant positive correlations, indicating that lower signal intensity and detectability are modestly associated with reduced correlation between P100 and EDTA, though additional factors likely also contribute. Spearman correlation coefficients and detectability values for all 127 biomarkers are provided in Supplementary Table 1.

**Figure 3.**
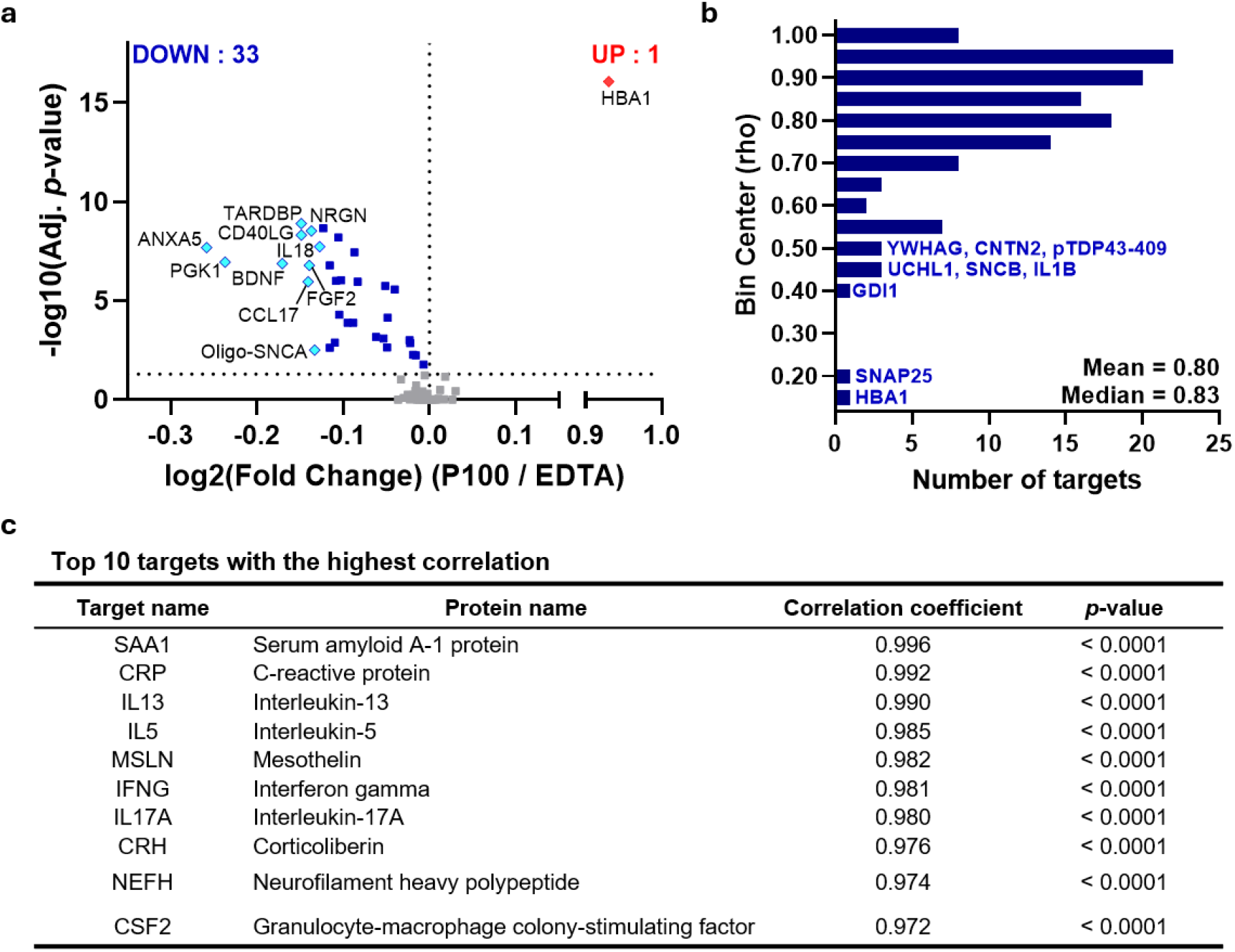
Comparison of biomarker levels between EDTA and P100. (a) Volcano plot comparing P100 and EDTA plasma biomarker concentrations. Thirty-three targets showed statistically significantly lower concentrations in P100 (log₂(FC) < 0). Light blue diamond dots indicate the top 10 targets with the largest fold changes; blue square dots indicate the remaining 23 targets with significantly lower concentrations in P100. One target, HBA1(red diamond dot), showed significantly higher concentration in P100 (log₂(FC) > 0). (b) Distribution of Spearman correlation coefficients between P100 and EDTA. Moderate to strong correlation (ρ ≥ 0.6) was observed for 111 targets, and strong correlation (ρ ≥ 0.8) was observed for 77 targets. For bin centers <0.55, biomarker target names are indicated next to the corresponding bars. (c) Top 10 targets with the highest correlation. HBA1, hemoglobin subunit alpha. Full names of all other protein targets are provided in the Abbreviations list and Supplementary Table 1.

### Concordance analysis for conventional AD biomarkers

We performed preliminary concordance analysis for eight conventional AD biomarkers and two biomarker ratios (Aβ42/Aβ40 and p-tau217/Aβ42) using both correlation and agreement methods (Figure 4). All eight biomarkers demonstrated statistically significant correlations (all *p* < 0.0001) between P100 and EDTA measurements. Six biomarkers showed strong correlation (ρ > 0.8), with NEFL achieving the highest correlation (ρ = 0.942). Aβ40 (ρ = 0.688) and p-tau217 (ρ = 0.570) showed moderate correlation. For biomarker ratios, p-tau217/Aβ42 showed strong correlation (ρ = 0.817, *p* < 0.0001), whereas Aβ42/Aβ40 showed weak correlation that did not reach statistical significance (ρ = 0.216, *p* = 0.075).

**Figure 4.**
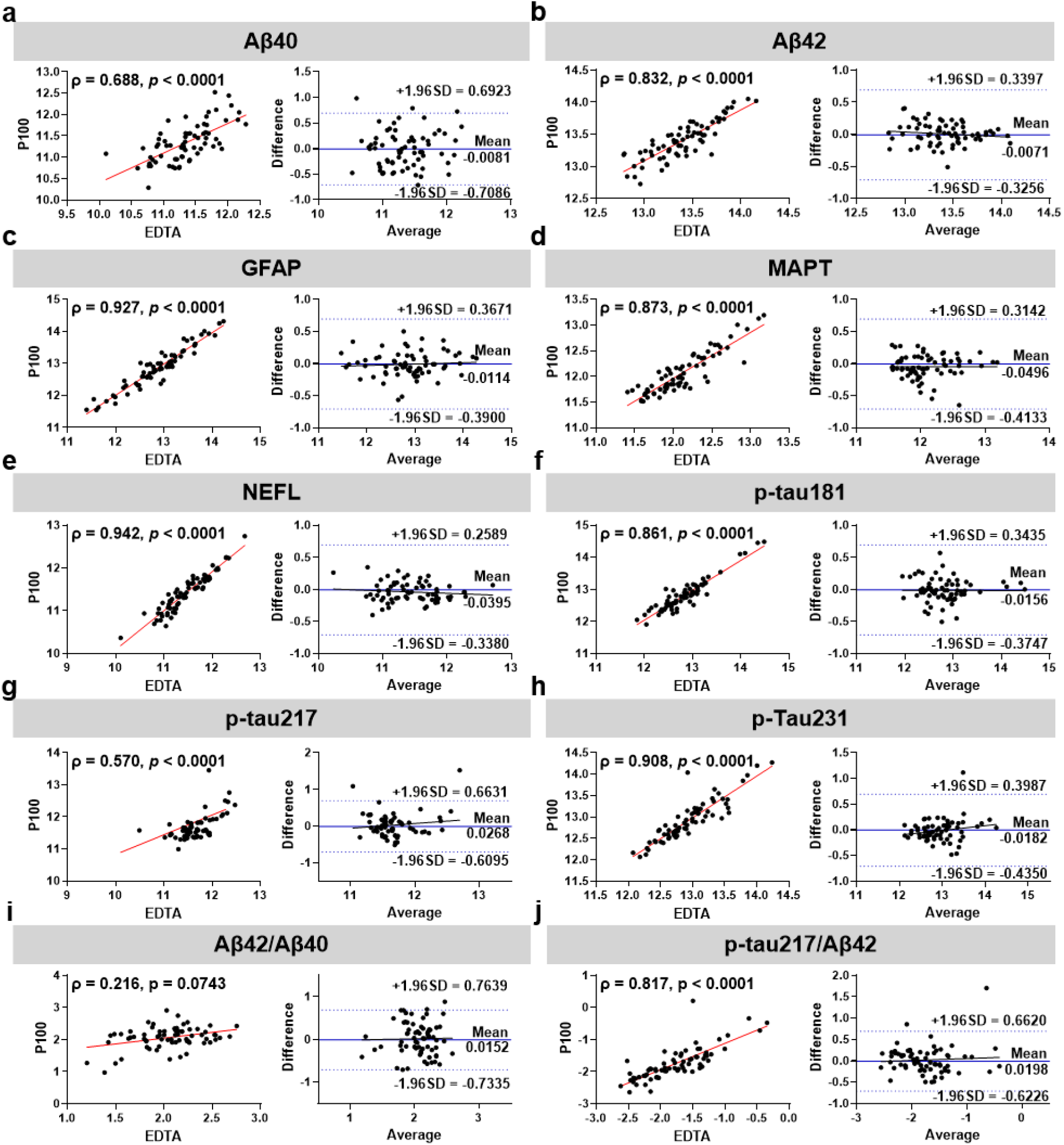
Correlation and agreement analysis of conventional AD biomarkers and ratios. (a-h) Correlation and Bland-Altman analysis for eight conventional AD biomarkers: Aβ40, Aβ42, GFAP, MAPT, NEFL, p-tau181, p-tau217, and p-tau231. (i-j) Correlation and agreement analysis for two biomarker ratios: Aβ42/Aβ40 (i) and p-tau217/Aβ42 (j). Each panel shows a correlation plot (left) and Bland-Altman plot (right). In correlation plots, each point represents a sample, with Spearman’s ρ and p-value indicated. The red solid line indicates the linear regression fit. In Bland-Altman plots, the x-axis shows the mean of P100 and EDTA measurements, and the y-axis shows the difference (P100 - EDTA). The solid line indicates mean bias, and dashed lines indicate ±1.96 standard deviation from the mean difference. Aβ40, amyloid beta 40; Aβ42, amyloid beta 42; GFAP, glial fibrillary acidic protein; MAPT, microtubule-associated protein tau; NEFL, neurofilament light polypeptide; p-tau181, tau phosphorylated at threonine 181; p-tau217, tau phosphorylated at threonine 217; p-tau231, tau phosphorylated at threonine 231.

Bland-Altman analysis was performed to assess systematic and proportional bias between P100 and EDTA measurements (Table 2, Figure 4). The mean differences (bias) were close to zero for all biomarkers (range: −0.050 to 0.027) and ratios (range: 0.015 to 0.020), indicating minimal systematic bias. Linear regression analysis revealed no significant proportional bias for any biomarker or ratio (all *p* > 0.05), indicating that the magnitude of differences was consistent across the measurement range. The distribution of differences for each biomarker is shown in Figure 4, with the proportion of points within ±1.96 SD ranging from 89.7% to 100% and within ±1 SD from 64.1% to 84.6% (Table 2). These findings demonstrate minimal systematic bias between P100 and EDTA plasma for conventional AD biomarkers.

**Table 2.**
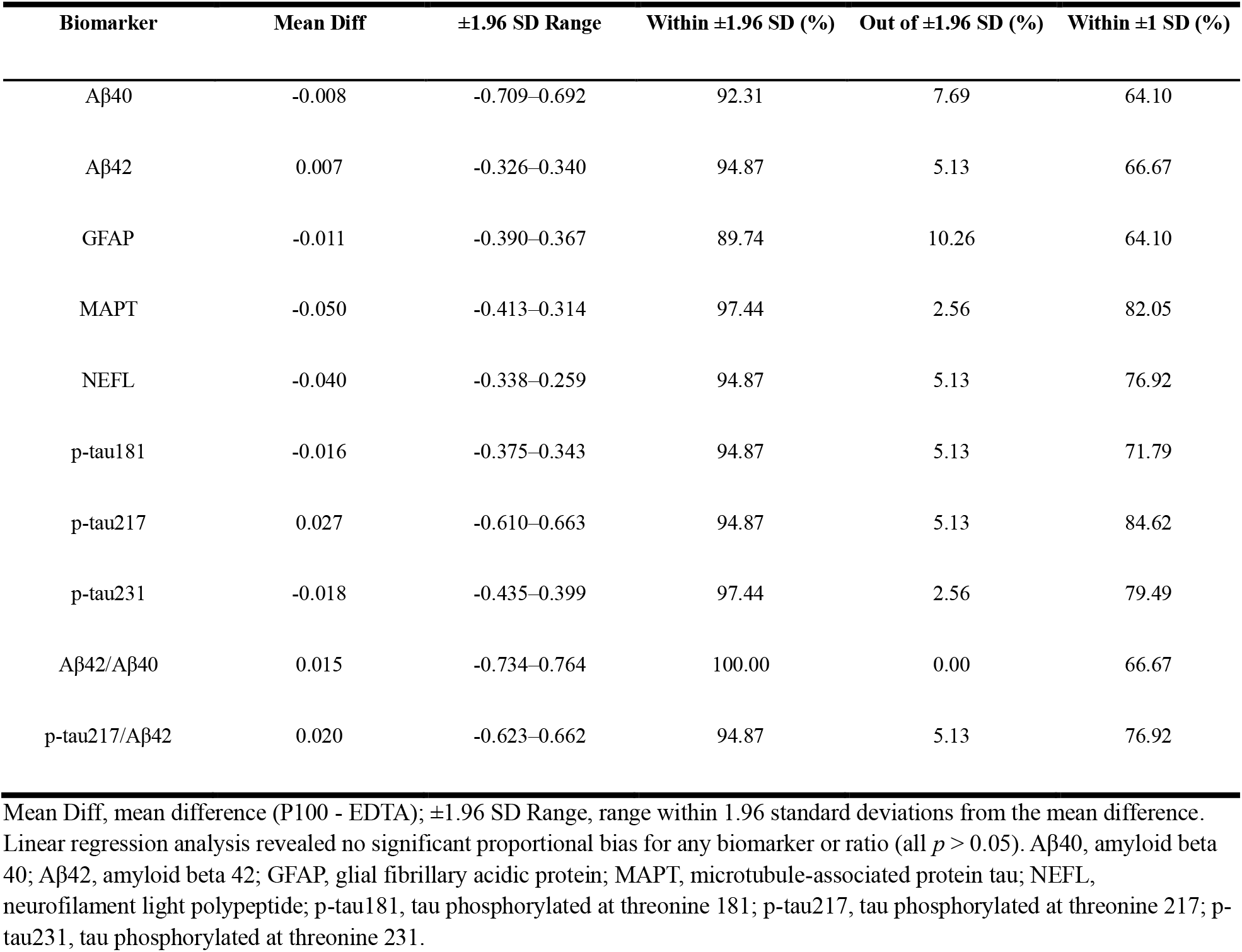
Bland-Altman analysis of conventional AD biomarkers and ratios.

| Biomarker | Mean Diff | $\pm 1.96$ SD Range | Within $\pm 1.96$ SD (%) | Out of $\pm 1.96$ SD (%) | Within $\pm 1$ SD (%) |
| --- | --- | --- | --- | --- | --- |
| Aβ40 | -0.008 | -0.709–0.692 | 92.31 | 7.69 | 64.10 |
| Aβ42 | 0.007 | -0.326–0.340 | 94.87 | 5.13 | 66.67 |
| GFAP | -0.011 | -0.390–0.367 | 89.74 | 10.26 | 64.10 |
| MAPT | -0.050 | -0.413–0.314 | 97.44 | 2.56 | 82.05 |
| NEFL | -0.040 | -0.338–0.259 | 94.87 | 5.13 | 76.92 |
| p-tau181 | -0.016 | -0.375–0.343 | 94.87 | 5.13 | 71.79 |
| p-tau217 | 0.027 | -0.610–0.663 | 94.87 | 5.13 | 84.62 |
| p-tau231 | -0.018 | -0.435–0.399 | 97.44 | 2.56 | 79.49 |
| Aβ42/Aβ40 | 0.015 | -0.734–0.764 | 100.00 | 0.00 | 66.67 |
| p-tau217/Aβ42 | 0.020 | -0.623–0.662 | 94.87 | 5.13 | 76.92 |

Mean Diff, mean difference (P100 - EDTA); $\pm 1.96$ SD Range, range within 1.96 standard deviations from the mean difference. Linear regression analysis revealed no significant proportional bias for any biomarker or ratio (all $p > 0.05$ ). Aβ40, amyloid beta 40; Aβ42, amyloid beta 42; GFAP, glial fibrillary acidic protein; MAPT, microtubule-associated protein tau; NEFL, neurofilament light polypeptide; p-tau181, tau phosphorylated at threonine 181; p-tau217, tau phosphorylated at threonine 217; p-tau231, tau phosphorylated at threonine 231.

To further evaluate concordance in amyloid PET status classification, we performed preliminary ROC analysis for differentiating Aβ PET status (Table 3, Figure 5). For the eight conventional AD biomarkers, four biomarkers—GFAP, p-tau181, tau phosphorylated at threonine 217 (p-tau217), and tau phosphorylated at threonine 231 (p-tau231)—demonstrated AUC values exceeding 0.7 with statistical significance (*p* < 0.05) in both P100 and EDTA plasma. Microtubule-associated protein tau (MAPT; measured as total tau) in EDTA and Aβ40 in P100 also showed AUC values above 0.7, but these did not reach statistical significance. Notably, p-tau217 showed identical AUC values (0.84) between the two tube types. For biomarker ratios, p-tau217/Aβ42 demonstrated high AUC values in both EDTA (0.79) and P100 (0.77), while the Aβ42/Aβ40 ratio showed divergent AUC values between P100 (0.84) and EDTA (0.55). DeLong test revealed no significant differences in AUC between EDTA and P100 for most biomarkers and ratios (*p* > 0.05), except for Aβ40 (*p* = 0.046) and Aβ42/Aβ40 ratio (*p* = 0.013) (Table 3).

**Figure 5.**
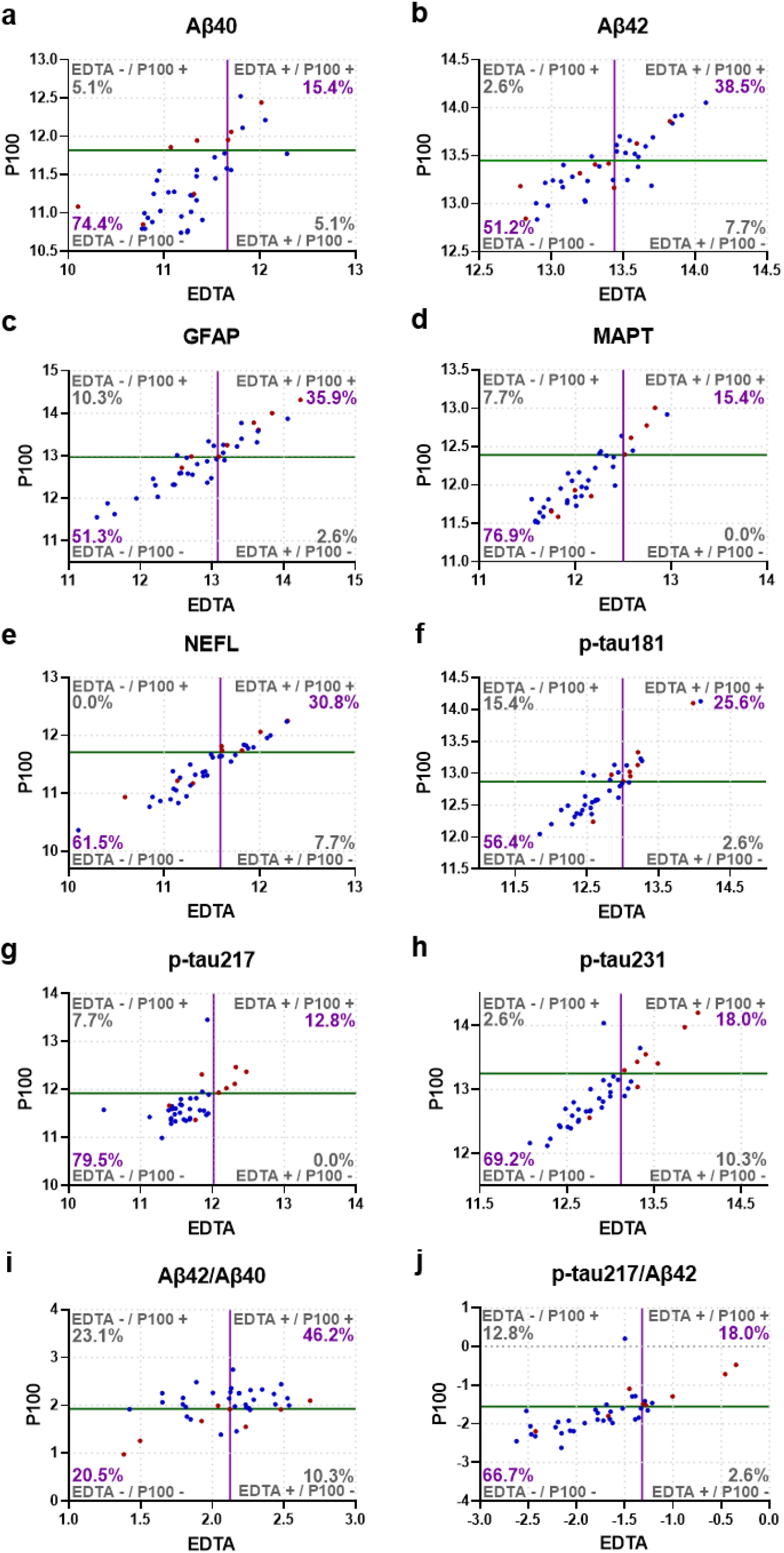
Preliminary amyloid PET status classification concordance using biomarker cutoff values. (a-h) Sample classification concordance for eight conventional AD biomarkers: Aβ40, Aβ42, GFAP, MAPT, NEFL, p-tau181, p-tau217, and p-tau231. (i-j) Sample classification concordance for two biomarker ratios: Aβ42/Aβ40 (i) and p-tau217/Aβ42 (j). ROC analysis was performed to evaluate performance in differentiating amyloid beta PET status (positive/negative), and optimal cutoff values were determined using Youden’s index. The purple vertical line indicates the EDTA cutoff, and the green horizontal line indicates the P100 cutoff. Each plot is divided into four quadrants representing classification outcomes: upper-right (both positive), lower-left (both negative), upper-left (EDTA negative/P100 positive), and lower-right (EDTA positive/P100 negative). Red dots indicate amyloid beta PET positive samples; blue dots indicate amyloid beta PET negative samples. Percentages in each quadrant indicate the proportion of samples. Concordant classifications (upper-right + lower-left) indicate agreement between EDTA and P100. Aβ40, amyloid beta 40; Aβ42, amyloid beta 42; GFAP, glial fibrillary acidic protein; MAPT, microtubule-associated protein tau; NEFL, neurofilament light polypeptide; p-tau181, tau phosphorylated at threonine 181; p-tau217, tau phosphorylated at threonine 217; p-tau231, tau phosphorylated at threonine 231.

**Table 3.** ROC analysis results for conventional AD biomarkers and ratios.

| | Aβ40 | Aβ42 | GFAP | MAPT | NEFL | p-tau181 | p-tau217 | p-tau231 | Aβ42/Aβ40 | p-tau217/Aβ42 |
| --- | --- | --- | --- | --- | --- | --- | --- | --- | --- | --- |
| **EDTA** |  |  |  |  |  |  |  |  |  |  |
| AUC | 0.53 | 0.60 | 0.77 | 0.69 | 0.60 | 0.82 | 0.84 | 0.91 | 0.55 | 0.79 |
| SE | 0.13 | 0.12 | 0.09 | 0.11 | 0.12 | 0.07 | 0.10 | 0.07 | 0.13 | 0.10 |
| 95% CI | 0.28–0.80 | 0.37–0.82 | 0.60–0.95 | 0.47–0.91 | 0.36–0.83 | 0.68–0.97 | 0.63–1.00 | 0.78–1.00 | 0.30–0.81 | 0.59–0.99 |
| P value | 0.781 | 0.404 | 0.02 | 0.102 | 0.404 | 0.005 | 0.004 | 0.000 | 0.651 | 0.012 |
| Cutoff | 11.66 | 13.44 | 13.08 | 12.50 | 11.59 | 13.00 | 12.02 | 13.12 | 2.16 | -1.32 |
| **P100** |  |  |  |  |  |  |  |  |  |  |
| AUC | 0.70 | 0.55 | 0.81 | 0.61 | 0.65 | 0.75 | 0.84 | 0.85 | 0.84 | 0.77 |
| SE | 0.11 | 0.12 | 0.08 | 0.13 | 0.12 | 0.11 | 0.10 | 0.09 | 0.07 | 0.10 |
| 95% CI | 0.48–0.92 | 0.32–0.78 | 0.65–0.96 | 0.36–0.86 | 0.41–0.88 | 0.54–0.95 | 0.64–1.00 | 0.68–1.00 | 0.70–0.98 | 0.58–0.97 |
| P value | 0.090 | 0.676 | 0.008 | 0.348 | 0.211 | 0.034 | 0.004 | 0.003 | 0.004 | 0.018 |
| Cutoff | 11.82 | 13.45 | 12.97 | 12.39 | 11.71 | 12.87 | 11.92 | 13.25 | 1.93 | -1.55 |
| **DeLong test** |  |  |  |  |  |  |  |  |  |  |
| P value | 0.046 | 0.466 | 0.409 | 0.067 | 0.164 | 0.216 | 1.000 | 0.131 | 0.013 | 0.759 |

ROC analysis was performed using amyloid beta PET status (positive/negative) as the reference standard. Optimal cutoff values were determined using Youden's index. DeLong test was used to compare AUC between EDTA and P100; no significant differences were observed (all $p > 0.05$ ) except for Aβ40 and Aβ42/Aβ40 ratio. AUC, area under the curve; SE, standard error; CI, confidence interval. Aβ40, amyloid beta 40; Aβ42, amyloid beta 42; GFAP, glial fibrillary acidic protein; MAPT, microtubule-associated protein tau; NEFL, neurofilament light polypeptide; p-tau181, tau phosphorylated at threonine 181; p-tau217, tau phosphorylated at threonine 217; p-tau231, tau phosphorylated at threonine 231.

Using Youden’s index-derived cutoff values, we assessed sample classification concordance between P100 and EDTA (Figure 5). Each scatter plot was divided into four quadrants by the cutoff values, where samples in the upper-right quadrant (positive in both tube types) and lower-left quadrant (negative in both tube types) represent concordant classification. The proportion of concordantly classified samples ranging from 82.1% (p-tau181) to 92.3% (MAPT, NEFL, and p-tau217) for individual biomarkers. For biomarker ratios, p-tau217/Aβ42 achieved 84.6% concordance, while Aβ42/Aβ40 showed 66.7%. These results on a preliminary cohort indicate that P100 and EDTA plasma demonstrate high agreement in classification for most conventional AD biomarkers.

## Discussion

In this study, we comprehensively evaluated the analytical performance of the P100 delayed centrifugation blood collection tubes relative to standard EDTA tubes across 127 neurodegeneration-related biomarkers using the NULISA platform. The NULISA assay – for both tube types - demonstrated excellent reproducibility, with mean and median coefficients of variation below 10% for both intra- and inter-plate measurements (Figure 1). Correlation analysis revealed strong concordance between P100 and EDTA plasma, with overall mean and median Spearman correlation coefficients exceeding 0.8 (Figure 3). Notably, eight conventional AD biomarkers showed robust agreement, with six biomarkers (NEFL, GFAP, p-tau231, MAPT, p-tqu181, and Aβ42) achieving correlation coefficients above 0.8 and minimal systematic bias with no proportional bias detected in Bland-Altman analysis (Figure 4). Two biomarker ratios were also evaluated: p-tau217/Aβ42 showed strong correlation (ρ = 0.817), whereas Aβ42/Aβ40 showed weak correlation (ρ = 0.216), though both showed minimal bias. Furthermore, to evaluate whether P100 and EDTA plasma yielded comparable classification performance, we performed ROC analysis using PET-assessed amyloid status as the reference standard. Both tube types showed comparable performance, with DeLong test confirming no significant AUC differences for most biomarkers. Sample classification concordance ranged from 82.1% to 92.3% across the eight conventional AD biomarkers, and 66.7%, 84.6% for biomarker ratios (Table 3, Figure 5). However, P100 samples exhibited slightly lower sample-level detectability in P100 than EDTA (94.7% vs. 95.9%; a difference of 1.17 percentage points) (Figure 2a, Supplementary Figure 1), and the majority of targets showed lower measured values in P100 compared to EDTA (Figure 3a).

The high reproducibility observed in our study, with CVs consistently below 10%, demonstrates the technical robustness of the NULISA assay (Figure 1a-d). This finding is particularly noteworthy given the large scale of our analysis encompassing 127 biomarkers using the NULISA platform, substantially expanding upon our previous work that evaluated six AD biomarkers^14^. This high-sensitivity multiplexed approach allowed simultaneous assessment of biomarkers related to amyloid and tau pathologies, neurodegeneration, and inflammation from minimal sample volumes. Together with prior cross-platform validation demonstrating strong concordance between NULISAseq measurements and established immunoassays for conventional AD biomarkers^17^, these findings support the technical reliability of the NULISA platform for large-scale biomarker studies. To our knowledge, this represents the first systematic analytical validation of P100 tubes using a high-sensitivity multiplex platform for an extensive panel of neurodegeneration-related biomarkers.

The strong correlation observed between P100 and EDTA measurements (mean and median Spearman ρ > 0.8) demonstrates that P100 tubes provide reliable quantification comparable to standard EDTA collection (Figure 3b). This concordance was particularly robust for the eight conventional AD biomarkers, with three achieving correlation coefficients exceeding 0.9 (NEFL, GFAP, and p-tau231) (Figure 4). Bland-Altman analysis further showed minimal systematic bias and no proportional bias for any biomarker (Table 2), indicating that the magnitude of differences between P100 and EDTA was consistent across the measurement range. These findings indicate that P100 can serve as a viable alternative to EDTA for neurodegeneration biomarker studies without introducing systematic bias.

While correlation and agreement analyses demonstrate quantitative concordance, the clinical utility of an alternative blood collection method ultimately depends on whether it yields consistent classification. Our ROC-based classification analysis addresses this by evaluating whether P100 and EDTA plasma categorize patients similarly using amyloid beta PET status as the reference standard. The high concordance rates observed (82.05%–92.31% for individual biomarkers, and 66.67%–84.62% for biomarker ratios) suggest that substituting P100 for EDTA would result in consistent patient classification for most cases. DeLong test showed no significant differences in AUC between EDTA and P100 for most biomarkers and ratios, except for Aβ40 (*p* = 0.046) and Aβ42/Aβ40 ratio (*p* = 0.013). Aβ peptides are subject to active proteolytic regulation by multiple endogenous proteases in vivo^12^. The differential AUC for Aβ40 may reflect the persistence of this proteolytic activity during blood collection and processing. Our previous work demonstrated that P100 tubes reduce Aβ peptide degradation compared to EDTA, while p-tau181, GFAP, and NEFL remain stable in both tube types^14^. The differential preservation of Aβ40 in P100 may therefore introduce modest shifts in absolute concentrations, contributing to the AUC differences observed for Aβ40 and the Aβ42/Aβ40 ratio. The lack of a corresponding effect on Aβ42 alone may reflect its lower plasma concentration relative to Aβ40.

Beyond these tube-specific effects, the overall classification performance of the Aβ42/Aβ40 ratio was limited and inconsistent across both tube types (AUC 0.55–0.84). This most likely reflects the low amyloid burden of this predominantly cognitively unimpaired, poor-sleep cohort: only 8 of 43 participants were amyloid-positive by visual read, and a number of these were near-threshold rather than unequivocally positive. Within such a narrow pathological range, ROC-based estimates for amyloid-tracking markers are inherently unstable and sensitive to the classification of individual cases. The between-tube difference observed for this ratio should therefore be interpreted with caution, as reflecting the limited amyloid signal in this cohort rather than superior performance of either tube type.

In contrast, p-tau217 illustrates how a low-abundance biomarker can still perform robustly for amyloid status classification. A previous study using the LUMIPULSE platform reported that plasma p-tau217 concentrations are approximately 10-fold lower than p-tau181^19^. Consistent with this, p-tau217 demonstrated reduced detectability in our cohort (80% in EDTA, 75% in P100; Supplementary Table 1) and only moderate correlation between tubes (ρ = 0.570). Despite these analytical limitations, p-tau217 achieved the highest AUC (0.84) and concordance rate (92.3%) among the eight conventional AD biomarkers in our ROC analysis. This aligns with previous reports demonstrating excellent performance of plasma p-tau217 for detecting amyloid positivity^20^. Together, these findings suggest that moderate correlation or analytical variability at low concentrations does not necessarily compromise diagnostic utility when sufficient separation exists between disease groups, and that consistency in status categorization across collection methods is particularly relevant for clinical trial screening or longitudinal monitoring.

Despite the strong overall concordance between P100 and EDTA, we observed modestly lower detectability in P100 samples (Figure 2a) and lower NPQ values in the majority of targets (Figure 3a). Volcano plot analysis revealed that 33 of 127 targets showed significantly lower concentrations in P100 compared to EDTA, while HBA1 was the only target with significantly higher concentration in P100. Nine targets showed weak correlation (ρ < 0.6) between P100 and EDTA (Figure 3b). Closer examination of these nine targets revealed two distinct patterns. Seven targets (UCHL1, SNCB, GDI1, pTDP43-409, IL1B, YWHAG, and SNAP25) ranked among the lowest 10% of all biomarkers in mean NPQ-LOD values, suggesting that proximity to detection limits likely contributes to their reduced correlation through increased measurement noise. In contrast, HBA1 and contactin-2 (CNTN2) showed sufficient detectability (ranks 88 and 91 of 127), indicating that other reasons may underlie their weak correlation. For HBA1, differential hemolysis between collection tubes is a likely contributor, as hemoglobin-related proteins are well-established markers of erythrocyte contamination in plasma proteomics^21^. The mechanism for CNTN2 remains unclear and warrants further investigation.

The elevation of HBA1 in P100 samples and the broader pattern of reduced detectability and lower NPQ values together suggest that differential hemolysis is a key contributor to the observed P100-EDTA differences. Hemolysis releases erythrocyte-derived proteases and peptidases into plasma, which can degrade some target analytes leading to decreased levels. This mechanism is consistent with both the elevation of erythrocyte-specific proteins (HBA1) and the simultaneous reduction in many other targets. Further mechanistic analysis is limited because the manufacturer does not currently disclose the composition of the proprietary protease inhibitor cocktail in P100 tubes. Without knowledge of which proteases are inhibited, it is not possible to predict which target proteins are protected from hemolysis-related degradation. Some prior evaluations have questioned the broad utility of P100 tubes; Aguilar-Mahecha et al. (2012)^10^ reported proteolytic protection for only a small subset of proteins, O’Neal et al. (2014) ^22^ reported modestly increased CVs for certain analytes in P100 compared to EDTA plasma in a COPD biomarker study. However, the context appears different for AD-specific biomarkers: our recent work demonstrated that P100 improves preservation of AD biomarkers at room temperature^14^.

Together with the strong P100-EDTA concordance observed across 127 neurodegeneration-related biomarkers in the present study, these findings support the continued consideration of P100 for AD biomarker research. These findings have several practical implications. The combination of P100’s room-temperature stability with reliable performance across a broad biomarker panel makes it particularly suitable for remote blood sampling in rural areas or regions with limited laboratory infrastructure, where samples may require extended transport times before processing. For multi-site clinical trials, the consistency of P100 measurements with EDTA reduces concerns about cross-site variability introduced by differential sample handling. Furthermore, the comprehensive validation across 127 neurodegeneration biomarkers extends the potential applications of P100 beyond AD to other neurodegenerative disease research and biomarker discovery efforts.

This study has several limitations. First, the cohort size was modest (*n* = 43), and few participants were deemed positive for amyloid based on PET (*n* = 8 positive, n=33 negative), which limits precision in classification concordance estimates. Second, although our findings demonstrate strong P100-EDTA concordance on the NULISA platform, generalization to other measurement platforms requires further validation. Future multi-site validation studies would help confirm and extend these findings.

In conclusion, the NULISA assay demonstrated high reproducibility for neurodegeneration biomarker measurement across an extensive panel of 127 targets. Using this platform, P100 blood collection tubes showed strong concordance with EDTA plasma, with minimal systematic bias and comparable classification performance for conventional AD biomarkers and biomarker ratios. By establishing that P100 tubes yield analytical and classification performance comparable to conventional EDTA tubes on the NULISA platform, these findings complement prior evidence for P100 biomarker stability and together support the use of P100 tubes for blood-based neurodegeneration biomarker research, including remote settings where immediate sample processing is not feasible.

## Acknowledgements

Thomas K. Karikari and the Karikari laboratory were supported by NIH/NIA (U01 AG098641, R01 AG083874, U24 AG082930, P30 AG066468, RF1 AG077474, R01 AG083156, R37 AG023651, R01 AG025516, R01 AG073267, R01 AG075336, R01 AG074971, U19 AG073153, U01 AG079828, P01 AG025204), NIH/NINDS (U01 NS131740, U01 NS141777, U24 NS141780), NIH/NIMH (R01 MH108509, R01 MH135096), Aging Mind Foundation (DAF2255207), DoD (HT94252320064), American Heart Association (24SFRNPCN1280228), Wellcome Leap (CARE-2025-7270262472), the Chuck Noll Foundation for Brain Injury Research, Greater Houston Community Foundation, Fujirebio, Genentech, and Astellas Pharma US. The ALPS study, Kristine A. Wilckens, and Rima F. Habte were supported by NIH/NIA grant R01AG068001. The content of this article is solely the responsibility of the authors and does not necessarily represent the official views of the funders.

## Author contributions

Y.L.: conceptualization, formal analysis, investigation, visualization, writing – original draft. X.Z.: conceptualization, methodology, visualization, writing – review & editing. M.F.F.: investigation, data curation. B.J.L.: investigation, formal analysis, writing – review & editing. R.F.H.: methodology, investigation, writing – review & editing. K.A.W.: resources, funding acquisition, writing – review & editing. T.K.K.: conceptualization, supervision, resources, funding acquisition, writing – review & editing.

## Data availability statement

The data supporting this study contain sensitive human participant information and cannot be deposited in a public repository due to institutional review board restrictions and the terms of participant consent. Specific de-identified data can be shared with qualified and identifiable investigators for the purpose of replicating the results and procedures in the study. Requests will be reviewed to ensure that data sharing requests conform to prevailing US legislation on data protection, intellectual property and confidentiality obligations. Establishment of a data transfer agreement may be necessary.

## Abbreviations

AD: Alzheimer’s disease;
ANXA5: annexin A5
APOE: apolipoprotein E
AUC: area under the curve
Aβ: amyloid beta
Aβ38/Aβ40/Aβ42: amyloid beta 38/40/42
BDNF: brain-derived neurotrophic factor
CCL17: C-C motif chemokine 17
CD40LG: CD40 ligand
CI: confidence interval
CNS: central nervous system
CNTN2: contactin-2
COPD: chronic obstructive pulmonary disease
CRH: corticoliberin
CRP: C-reactive protein
CSF: cerebrospinal fluid
CSF2: granulocyte-macrophage colony-stimulating factor
CV: coefficient of variation
CXCL10: C-X-C motif chemokine 10
EDTA: ethylenediaminetetraacetic acid
FC: fold change
FDR: false discovery rate
FGF2: fibroblast growth factor 2
FLT1: vascular endothelial growth factor receptor 1
GDI1: Rab GDP dissociation inhibitor alpha
GDNF: glial cell line-derived neurotrophic factor
GFAP: glial fibrillary acidic protein
HBA1: hemoglobin subunit alpha
IFNG: interferon gamma
IL1B: interleukin-1 beta
IL5: interleukin-5
IL13: interleukin-13
IL17A: interleukin-17A
IL18: interleukin-18
LOD: limit of detection
MAPT: microtubule-associated protein tau
MME: membrane metalloendopeptidase
MSLN: mesothelin
NEFH: neurofilament heavy polypeptide
NEFL: neurofilament light polypeptide (also abbreviated NfL)
NPQ: NULISA Protein Quantification
NPTX1: neuronal pentraxin-1
NPTXR: neuronal pentraxin receptor
NRGN: neurogranin
NULISA: NUcleic acid-Linked Immuno-Sandwich Assay
Oligo-SNCA: oligomeric alpha-synuclein
PDLIM5: PDZ and LIM domain 5
PET: positron emission tomography
PGK1: phosphoglycerate kinase 1
PiB: Pittsburgh Compound B
PSEN1: presenilin 1
PTN: pleiotrophin
p-tau: phosphorylated tau
pTDP43-409: TAR DNA-binding protein 43 phosphorylated at serine 409
ROC: receiver operating characteristic
S100A12: protein S100-A12
S100B: S100 calcium binding protein B
SAA1: serum amyloid A-1 protein
SD: standard deviation
SE: standard error
SFTPD: pulmonary surfactant-associated protein D
SNAP25: synaptosomal-associated protein 25
SNCA: alpha-synuclein
SNCB: synuclein beta
SOD1: superoxide dismutase [Cu-Zn]
SQSTM1: sequestosome-1
TARDBP: TAR DNA-binding protein 43
YWHAG: tyrosine 3-monooxygenase/tryptophan 5-monooxygenase activation protein gamma
YWHAZ: 14-3-3 protein zeta/delta

