## Supplementary Table 1 for "Analytical performance of a delayed-centrifugation blood collection tube for remote blood biomarker assessment"

| Index | Target Name | Protein Name | Correlation rho_tube | Correlation P-value_tube | Detectability P100 | Detectability EDTA |
|---|---|---|---|---|---|---|
| 1 | ACHE | Acetylcholinesterase | 0.82 | < 0.0001 | 100.0 | 100.0 |
| 2 | AGRN | Agrin | 0.78 | < 0.0001 | 100.0 | 100.0 |
| 3 | ANXA5 | Annexin A5 | 0.81 | < 0.0001 | 100.0 | 100.0 |
| 4 | APOE | Apolipoprotein E | 0.93 | < 0.0001 | 100.0 | 100.0 |
| 5 | APOE4 | Apolipoprotein E | 0.70 | < 0.0001 | 100.0 | 100.0 |
| 6 | ARSA | Arylsulfatase A | 0.75 | < 0.0001 | 100.0 | 100.0 |
| 7 | Aβ38 | Amyloid-beta precursor protein 38 | 0.54 | < 0.0001 | 100.0 | 100.0 |
| 8 | Aβ40 | Amyloid-beta precursor protein 40 | 0.69 | < 0.0001 | 100.0 | 100.0 |
| 9 | Aβ42 | Amyloid-beta precursor protein 42 | 0.83 | < 0.0001 | 100.0 | 100.0 |
| 10 | BACE1 | Beta-secretase 1 | 0.81 | < 0.0001 | 100.0 | 100.0 |
| 11 | BASP1 | Brain abundant membrane attached signal protein 1 | 0.93 | < 0.0001 | 93.3 | 100.0 |
| 12 | BDNF | Brain-derived neurotrophic factor | 0.74 | < 0.0001 | 100.0 | 100.0 |
| 13 | CALB2 | Calretinin | 0.82 | < 0.0001 | 100.0 | 100.0 |
| 14 | CCL11 | Eotaxin | 0.94 | < 0.0001 | 100.0 | 100.0 |
| 15 | CCL13 | C-C motif chemokine 13 | 0.73 | < 0.0001 | 100.0 | 100.0 |
| 16 | CCL17 | C-C motif chemokine 17 | 0.82 | < 0.0001 | 100.0 | 100.0 |
| 17 | CCL2 | C-C motif chemokine 2 | 0.80 | < 0.0001 | 100.0 | 100.0 |
| 18 | CCL22 | C-C motif chemokine 22 | 0.90 | < 0.0001 | 100.0 | 100.0 |
| 19 | CCL26 | C-C motif chemokine 26 | 0.89 | < 0.0001 | 100.0 | 100.0 |
| 20 | CCL3 | C-C motif chemokine 3 | 0.85 | < 0.0001 | 100.0 | 100.0 |
| 21 | CCL4 | C-C motif chemokine ligand 4 | 0.91 | < 0.0001 | 100.0 | 100.0 |
| 22 | CD40LG | CD40 ligand | 0.76 | < 0.0001 | 100.0 | 100.0 |
| 23 | CD63 | CD63 antigen | 0.95 | < 0.0001 | 100.0 | 100.0 |
| 24 | CHI3L1 | Chitinase-3-like protein 1 | 0.96 | < 0.0001 | 100.0 | 100.0 |
| 25 | CHIT1 | Chitotriosidase-1 | 0.97 | < 0.0001 | 100.0 | 100.0 |
| 26 | CNTN2 | Contactin-2 | 0.51 | < 0.0001 | 100.0 | 100.0 |
| 27 | CRH | Corticoliberin | 0.98 | < 0.0001 | 100.0 | 100.0 |
| 28 | CRP | C-reactive protein | 0.99 | < 0.0001 | 100.0 | 100.0 |
| 29 | CSF2 | Granulocyte-macrophage colony-stimulating factor | 0.97 | < 0.0001 | 100.0 | 100.0 |
| 30 | CST3 | Cystatin-C | 0.84 | < 0.0001 | 100.0 | 100.0 |
| 31 | CX3CL1 | Fractalkine | 0.79 | < 0.0001 | 100.0 | 100.0 |
| 32 | CXCL1 | Growth-regulated alpha protein | 0.86 | < 0.0001 | 100.0 | 100.0 |
| 33 | CXCL10 | C-X-C motif chemokine 10 | 0.88 | < 0.0001 | 100.0 | 100.0 |
| 34 | CXCL8 | Interleukin-8, IL8 | 0.68 | < 0.0001 | 100.0 | 100.0 |
| 35 | DDC | NA | 0.96 | < 0.0001 | 100.0 | 100.0 |
| 36 | ENO2 | Gamma-enolase | 0.77 | < 0.0001 | 100.0 | 100.0 |
| 37 | FABP3 | Fatty acid-binding protein, heart | 0.94 | < 0.0001 | 100.0 | 100.0 |
| 38 | FCN2 | Ficolin-2 | 0.89 | < 0.0001 | 100.0 | 100.0 |
| 39 | FGF2 | Fibroblast growth factor 2 | 0.86 | < 0.0001 | 100.0 | 100.0 |
| 40 | FLT1 | Vascular endothelial growth factor receptor 1 | 0.65 | < 0.0001 | 100.0 | 100.0 |
| 41 | FOLR1 | Folate receptor alpha | 0.78 | < 0.0001 | 100.0 | 100.0 |
| 42 | GDF15 | Growth/differentiation factor 15 | 0.96 | < 0.0001 | 100.0 | 100.0 |
| 43 | GDI1 | Rab GDP dissociation inhibitor alpha | 0.40 | < 0.001 | 54.7 | 73.2 |
| 44 | GDNF | Glial cell line-derived neurotrophic factor | 0.95 | < 0.0001 | 76.0 | 73.2 |
| 45 | GFAP | Glial fibrillary acidic protein | 0.93 | < 0.0001 | 100.0 | 100.0 |
| 46 | GOT1 | Aspartate aminotransferase, cytoplasmic | 0.57 | < 0.0001 | 100.0 | 100.0 |
| 47 | HBA1 | Hemoglobin subunit alpha | 0.15 | 0.233 | 100.0 | 100.0 |
| 48 | HTT | Huntingtin | 0.77 | < 0.0001 | 100.0 | 100.0 |
| 49 | ICAM1 | Intercellular adhesion molecule 1 | 0.87 | < 0.0001 | 100.0 | 100.0 |
| 50 | IFNG | Interferon gamma | 0.98 | < 0.0001 | 100.0 | 100.0 |
| 51 | IGF1R | Insulin-like growth factor 1 receptor | 0.53 | < 0.0001 | 100.0 | 100.0 |
| 52 | IGFBP7 | Insulin-like growth factor-binding protein 7 | 0.71 | < 0.0001 | 100.0 | 100.0 |
| 53 | IL10 | Interleukin-10 | 0.95 | < 0.0001 | 100.0 | 100.0 |
| 54 | IL12p70 | Interleukin-12 subunit beta\|Interleukin-12 subunit alpha | 0.94 | < 0.0001 | 100.0 | 100.0 |
| 55 | IL13 | Interleukin-13 | 0.99 | < 0.0001 | 94.7 | 94.4 |
| 56 | IL15 | Interleukin-15 | 0.88 | < 0.0001 | 100.0 | 100.0 |
| 57 | IL16 | Pro-interleukin-16 | 0.83 | < 0.0001 | 100.0 | 100.0 |
| 58 | IL17A | Interleukin-17A | 0.98 | < 0.0001 | 100.0 | 97.2 |
| 59 | IL18 | Interleukin-18 | 0.86 | < 0.0001 | 100.0 | 100.0 |
| 60 | IL1B | Interleukin-1 beta | 0.47 | < 0.0001 | 49.3 | 77.5 |
| 61 | IL2 | Interleukin-2 | 0.87 | < 0.0001 | 100.0 | 100.0 |
| 62 | IL33 | Interleukin 33 | 0.90 | < 0.0001 | 100.0 | 100.0 |
| 63 | IL4 | Interleukin-4 | 0.69 | < 0.0001 | 100.0 | 94.4 |
| 64 | IL5 | Interleukin-5 | 0.98 | < 0.0001 | 100.0 | 100.0 |
| 65 | IL6 | Interleukin-6 | 0.95 | < 0.0001 | 100.0 | 100.0 |
| 66 | IL6R | Interleukin-6 receptor subunit alpha | 0.75 | < 0.0001 | 100.0 | 100.0 |
| 67 | IL7 | Interleukin-7 | 0.62 | < 0.0001 | 100.0 | 100.0 |
| 68 | IL9 | Interleukin-9 | 0.91 | < 0.0001 | 100.0 | 100.0 |
| 69 | KDR | Vascular endothelial growth factor receptor 2 | 0.92 | < 0.0001 | 100.0 | 100.0 |
| 70 | KLK6 | Kallikrein-6 | 0.85 | < 0.0001 | 100.0 | 100.0 |
| 71 | MAPT | Microtubule-associated protein tau; total tau | 0.87 | < 0.0001 | 100.0 | 100.0 |

| | | | | | | |
|---|---|---|---|---|---|---|
| 72 | MDH1 | Malate dehydrogenase, cytoplasmic | 0.68 | < 0.0001 | 100.0 | 100.0 |
| 73 | MME | Membrane metalloendopeptidase | 0.96 | < 0.0001 | 100.0 | 100.0 |
| 74 | MSLN | Mesothelin | 0.98 | < 0.0001 | 100.0 | 100.0 |
| 75 | NEFH | Neurofilament heavy polypeptide | 0.97 | < 0.0001 | 100.0 | 100.0 |
| 76 | NEFL | Neurofilament light polypeptide | 0.94 | < 0.0001 | 100.0 | 100.0 |
| 77 | NGF | Beta-nerve growth factor | 0.87 | < 0.0001 | 100.0 | 100.0 |
| 78 | NPTX1 | Neuronal pentraxin-1 | 0.73 | < 0.0001 | 100.0 | 100.0 |
| 79 | NPTX2 | Neuronal pentraxin-2 | 0.88 | < 0.0001 | 100.0 | 100.0 |
| 80 | NPTXR | Neuronal pentraxin receptor | 0.71 | < 0.0001 | 100.0 | 100.0 |
| 81 | NPY | Neuropeptide Y | 0.86 | < 0.0001 | 100.0 | 100.0 |
| 82 | NRGN | Neurogranin | 0.75 | < 0.0001 | 100.0 | 100.0 |
| 83 | Oligo-SNCA | Oligomeric alpha-synuclein | 0.76 | < 0.0001 | 100.0 | 95.8 |
| 84 | PARK7 | Protein/nucleic acid deglycase DJ-1 | 0.82 | < 0.0001 | 100.0 | 100.0 |
| 85 | PDGFRB | Platelet-derived growth factor receptor beta | 0.92 | < 0.0001 | 100.0 | 100.0 |
| 86 | PDLIM5 | PDZ and LIM domain 5 | 0.95 | < 0.0001 | 86.7 | 90.1 |
| 87 | PGF | Placenta growth factor | 0.73 | < 0.0001 | 100.0 | 100.0 |
| 88 | PGK1 | Phosphoglycerate kinase 1 | 0.81 | < 0.0001 | 100.0 | 100.0 |
| 89 | POSTN | Periostin | 0.91 | < 0.0001 | 100.0 | 100.0 |
| 90 | PRDX6 | Peroxiredoxin-6 | 0.81 | < 0.0001 | 100.0 | 100.0 |
| 91 | PSEN1 | Presenilin 1 | 0.66 | < 0.0001 | 100.0 | 100.0 |
| 92 | pSNCA-129 | Alpha-synuclein | 0.81 | < 0.0001 | 100.0 | 100.0 |
| 93 | pTau-181 | Tau phosphorylated at threonine 181 | 0.86 | < 0.0001 | 100.0 | 100.0 |
| 94 | pTau-217 | Tau phosphorylated at threonine 217 | 0.57 | < 0.0001 | 74.7 | 78.9 |
| 95 | pTau-231 | Tau phosphorylated at threonine 231 | 0.91 | < 0.0001 | 100.0 | 100.0 |
| 96 | pTDP43-409 | TAR DNA-binding protein 43 phosphorylated at serine 409 | 0.52 | < 0.0001 | 32.0 | 67.6 |
| 97 | PTN | Pleiotrophin | 0.54 | < 0.0001 | 54.7 | 66.2 |
| 98 | REST | RE1 silencing transcription factor | 0.88 | < 0.0001 | 100.0 | 100.0 |
| 99 | RUVBL2 | RuvB like AAA ATPase 2 | 0.67 | < 0.0001 | 100.0 | 100.0 |
| 100 | S100A12 | Protein S100-A12 | 0.77 | < 0.0001 | 100.0 | 100.0 |
| 101 | S100B | S100 calcium binding protein B | 0.75 | < 0.0001 | 100.0 | 100.0 |
| 102 | SAA1 | Serum amyloid A-1 protein | 1.00 | < 0.0001 | 100.0 | 100.0 |
| 103 | SFRP1 | Secreted frizzled-related protein 1 | 0.62 | < 0.0001 | 100.0 | 100.0 |
| 104 | SFTPD | Pulmonary surfactant-associated protein D | 0.95 | < 0.0001 | 100.0 | 100.0 |
| 105 | SLIT2 | Slit homolog 2 protein | 0.89 | < 0.0001 | 100.0 | 100.0 |
| 106 | SMOC1 | SPARC-related modular calcium-binding protein 1 | 0.94 | < 0.0001 | 100.0 | 100.0 |
| 107 | SNAP25 | Synaptosomal-associated protein 25 | 0.21 | 0.087 | 100.0 | 100.0 |
| 108 | SNCA | Alpha-synuclein | 0.74 | < 0.0001 | 100.0 | 100.0 |
| 109 | SNCB | Synuclein beta | 0.46 | < 0.0001 | 9.3 | 40.8 |
| 110 | SOD1 | Superoxide dismutase [Cu-Zn] | 0.57 | < 0.0001 | 100.0 | 100.0 |
| 111 | SQSTM1 | Sequestosome-1 | 0.79 | < 0.0001 | 100.0 | 100.0 |
| 112 | TAFA5 | Chemokine-like protein TAFA-5 | 0.79 | < 0.0001 | 100.0 | 100.0 |
| 113 | TARDBP | TAR DNA-binding protein 43 | 0.81 | < 0.0001 | 100.0 | 100.0 |
| 114 | TEK | Angiopoietin-1 receptor | 0.87 | < 0.0001 | 100.0 | 100.0 |
| 115 | TIMP3 | Metalloproteinase inhibitor 3 | 0.75 | < 0.0001 | 100.0 | 100.0 |
| 116 | TNF | Tumor necrosis factor | 0.92 | < 0.0001 | 100.0 | 100.0 |
| 117 | TREM1 | Triggering receptor expressed on myeloid cells 1 | 0.90 | < 0.0001 | 100.0 | 100.0 |
| 118 | TREM2 | Triggering receptor expressed on myeloid cells 2 | 0.95 | < 0.0001 | 100.0 | 100.0 |
| 119 | UBB | Polyubiquitin-B | 0.78 | < 0.0001 | 32.0 | 42.3 |
| 120 | UCHL1 | Ubiquitin carboxyl-terminal hydrolase isozyme L1 | 0.45 | < 0.001 | 0.0 | 0.0 |
| 121 | VCAM1 | Vascular cell adhesion protein 1 | 0.84 | < 0.0001 | 100.0 | 100.0 |
| 122 | VEGFA | Vascular endothelial growth factor A | 0.78 | < 0.0001 | 100.0 | 100.0 |
| 123 | VEGFD | Vascular endothelial growth factor D | 0.83 | < 0.0001 | 100.0 | 98.6 |
| 124 | VGF | VGF nerve growth factor inducible | 0.92 | < 0.0001 | 97.3 | 93.0 |
| 125 | VSNL1 | Visinin-like protein 1 | 0.90 | < 0.0001 | 100.0 | 100.0 |
| 126 | YWHAG | Tyrosine 3-monooxygenase/tryptophan 5-monooxygenase activation protein gamma | 0.48 | < 0.0001 | 96.0 | 100.0 |
| 127 | YWHAZ | 14-3-3 protein zeta/delta | 0.57 | < 0.0001 | 28.0 | 57.7 |
