## Supplementary Figure 1 for "Analytical performance of a delayed-centrifugation blood collection tube for remote blood biomarker assessment"

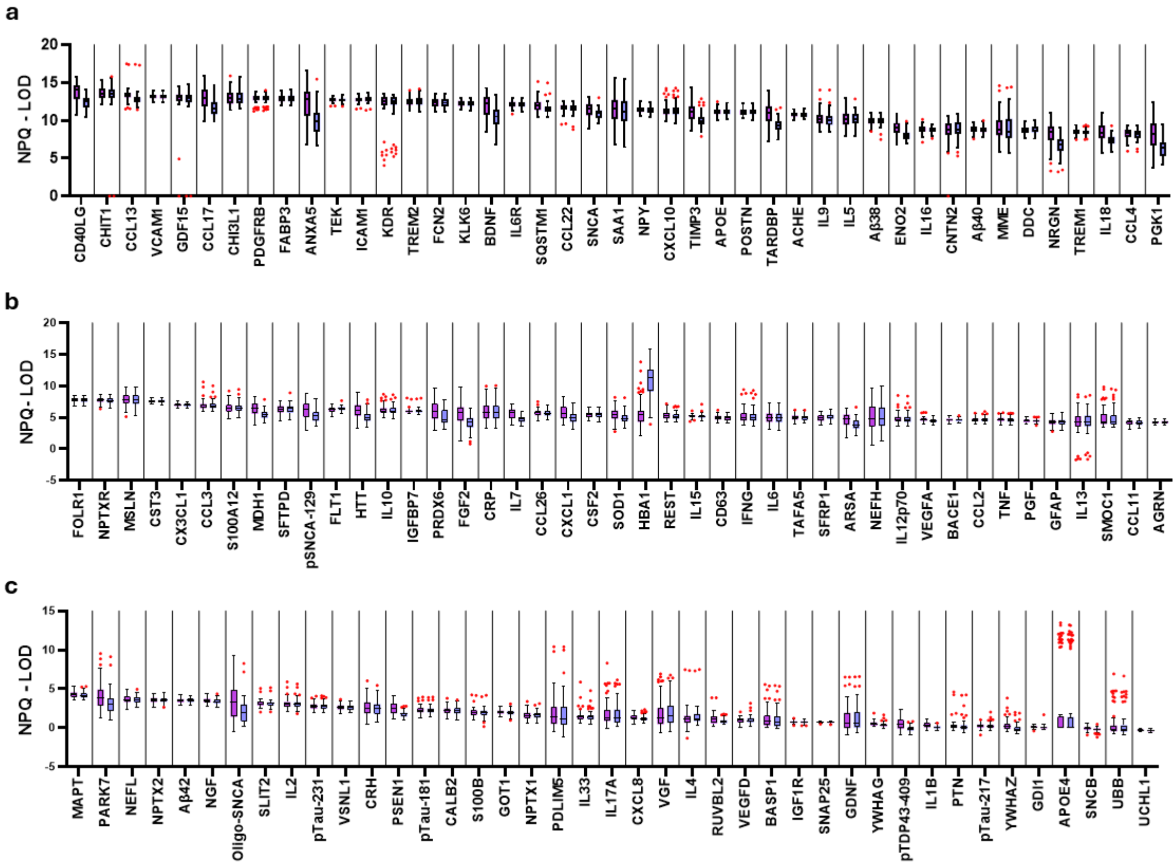


**Supplementary Fig 1. Measurement levels above detection limit by target in P100 and EDTA**

(a-c) Detectability box plots for each of the 127 biomarker targets in EDTA (left, purple) and P100 (right, blue) groups. The x-axis displays target names, and the y-axis shows NULISA protein quantification (NPQ) minus the limit of detection (LOD). Higher values indicate greater detectability above the detection threshold. Targets are arranged in descending order by median NPQ-LOD value: (a) top 43 targets, (b) next 43 targets, and (c) remaining 41 targets. Full names of all other protein targets are provided in the Abbreviations list and Supplementary Table 1.
